# Association Between the Use of Cholesterol-Lowering Prescription Medications and Gastric Cancer (GC): An Analysis from the NHANES Database

**DOI:** 10.1101/2025.05.20.25327984

**Authors:** Peng Dai, Jialin Zhou

**Affiliations:** General Medicine Department, Shanxi Province Cancer Hospital/Shanxi Hospital Affiliated to Cancer Hospital, Chinese Academy of Medical Sciences/Cancer Hospital Affiliated to Shanxi Medical University, Taiyuan, Shanxi, 030013, PRC China; Wuhan Vocational College of Software and Engineering (Wuhan Open University), Wuhan, Hubei, 430205, PRC China

**Keywords:** Gastric cancer, use of cholesterol-lowering prescription medications, NHANES, Risk stratification, ROC

## Abstract

The relationship between Gastric cancer (GC) and various risk factors is well-established, but the relationship between use of cholesterol-lowering prescription medications and GC remains controversial. This study aimed to investigate further the association between the two utilizing the National Health and Nutrition Examination Survey (NHANES) database. In this study, the NHANES database (1999-2018) was utilized to explore the association between the use of cholesterol-lowering prescription medications and GC. Baseline characteristics were first outlined using data from NHANES, followed by association analysis, risk stratification analysis, and receiver operating characteristic (ROC) analysis. Baseline statistical results indicated that the use of cholesterol-lowering prescription medications was significantly associated with GC, both as an independent exposure factor and after adjusting for various covariates. The results of risk stratification analysis showed a significant positive correlation between the use of cholesterol-lowering prescription medications and GC (odds ratio (OR) = 1.73, 95% confidence intervals (CI) = 1.07-2.8, p < 0.05). Additionally, ROC analysis result indicated that use of cholesterol-lowering prescription medications was a risk predictor for GC and could relatively accurately distinguish between GC patients and non-GC patients (AUC = 0.799). GC was associated with cholesterol, and use of cholesterol-lowering prescription medications was a risk predictor for GC. This provides valuable insights for future research.

## 1 Introduction

Gastric cancer (GC) is the fifth most common malignancy worldwide and the third leading cause of cancer-related mortality. Key risk factors for GC include Helicobacter pylori infection, advanced age, high salt intake, and diets low in fruits and vegetable(1). The pathogenesis of GC is recognized as a multifactorial process, entailing intricate interplay among genetic susceptibility, environmental exposures, and modifiable lifestyle determinants.. Interestingly, recent studies have identified an inverse relationship between total cholesterol levels and GC risk. Cholesterol, a fundamental component of cellular membranes, is essential for maintaining cellular integrity and function(2). Dysregulation of cholesterol metabolism may promote cancer cell proliferation and metastasis.

Several mechanisms have been proposed to explain how elevated cholesterol levels may enhance the development of gastric cancer, potentially through altering cell membrane structures, amplifying oncogenic signaling pathways, or fostering a pro-inflammatory microenvironment. A systematic review conducted by Lu et al. demonstrated a significant positive correlation between elevated serum total cholesterol levels and the prevalence of GI cancers. Furthermore, dietary patterns characterized by high fat content coupled with deficient intake of complex carbohydrates, fresh fruits, and vegetables have been identified as a significant risk factor for gastric cancer GC development(3). A meta-analysis supported by Hu J, Current evidence suggests that dietary patterns characterized by elevated saturated fat intake are associated with an increased risk of gastric adenocarcinoma, particularly in individuals with obesity and those presenting with malignancies localized to the gastric cardia region(4). Additionally, the differential effects of various cholesterol-lowering agents on GC risk have yet to be conclusively established. Clarifying these associations is of significant clinical importance, as it could inform more tailored therapeutic strategies and improve patient outcomes(5).

The National Health and Nutrition Examination Survey (NHANES), a comprehensive research program administered by the National Center for Health Statistics (NCHS) under the U.S. Centers for Disease Control and Prevention (CDC), provides valuable data for investigating these associations. NHANES collects extensive information on demographics, health conditions, dietary habits, and biomarkers through questionnaires, physical examinations, and laboratory tests. Since 1999, NHANES has been conducted continuously, offering a robust dataset for examining the prevalence of diagnosed and undiagnosed conditions, nutrient intake, and chemical exposures(6).

This study utilizes NHANES data to explore the potential association between cholesterol-lowering prescription medications and GC risk. As the interplay between cholesterol metabolism and GC pathogenesis garners increasing research attention, this investigation aims to provide novel insights into the role of cholesterol-lowering drugs, particularly statins, in GC development.

## 2 Materials and methods

### 2.1 Data collection

All participants were from the National Health and Nutrition Examination Survey (NHANES) database, and provided written informed consent. Studies that have completed data collection. Participants aged 20 years and above were included, and had been explicitly informed by a doctor or health expert about their GC status or were aware of not having any malignant tumors; and completeness of primary study variables. Exclusion criteria were refusal to answer questions related to GC diagnosis, responses of “unknown”, or missing data in this section; and missing primary study variables. This analysis finally included 51,630 non-GC individuals and 36 GC patients from 1999 to 2018 (**Figure 1**).

**Figure 1.**
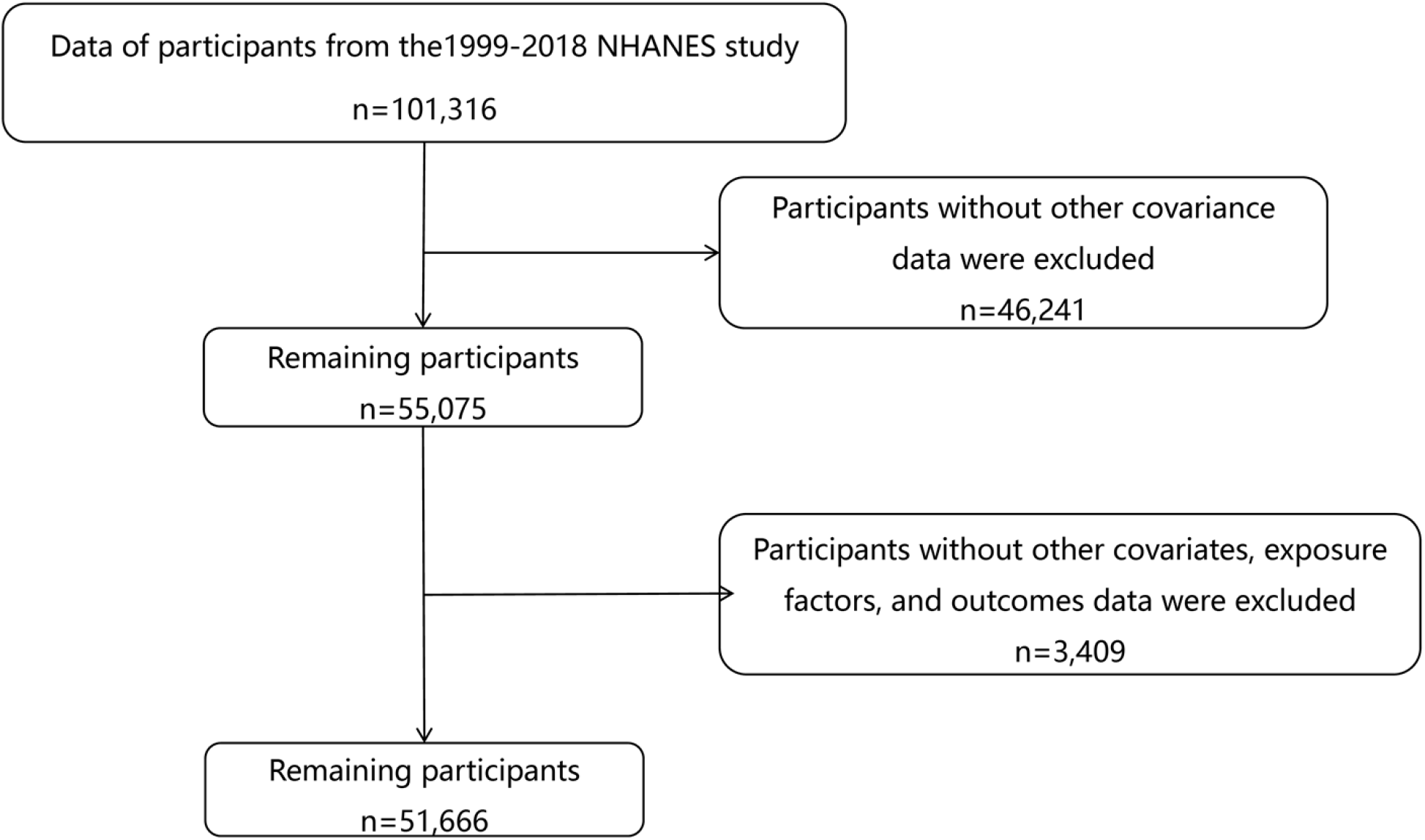
Study population selection process for Gastric Cancer (GC) analysis in NHANES (1999-2018).

### 2.2 NHANES variables

To better evaluate NHANES data, this study defined variables. Participants who were previously informed by a doctor or health expert that they had cancer or malignancy and answered “yes” were classified as GC cases if they also responded “stomach” to the question, which asked about the type of cancer. Participants who answered “no” to the question were classified as the healthy control group. The exposure factor was applied to subjects who had used cholesterol-lowering prescription drugs. Several important covariates (age, gender, education (%), race, family income-to-poverty ratio (PIR), smoke (%), hypertension, body mass index (BMI), diabetes, systemic inflammation index (SII)) were selected to assess the impact of potential confounding factors

### 2.3 Description of baseline characteristics of subjects

Subsequently, the baseline characteristics were described, with categorical variables expressed as percentages. Categorical variables and continuous variables were separately analyzed using the weighted chi-square test and t-tests.

### 2.4 Risk correlation analysis

To identify exposure factors significantly associated with GC, the “nhanesA” (v 1.0.3) (7) packages **was** utilized to construct 3 weighted multivariate logistic regression models. The odds ratio (OR) and 95% confidence intervals (CI) were then calculated. Based on the confounders, the NHANES disease models were defined as follows: model 1 was unadjusted; model 2 was based on model 1 and adjusted for age, race, and gender, serving as the minimally adjusted model; model 3 was based on model 2 and further adjusted for educational level, PIR, BMI, smoke, hypertension, diabetes, and SII, serving as the fully adjusted model.

### 2.5 Risk stratification analysis based on model 3

To further confirm the stability of the correlation between exposure factors and outcome risk, the correlation between covariates and outcome variables was analyzed using weighted logistic regression. The results were then visualized using a forest plot.

### 2.6 Receiver operating characteristic (ROC) analysis

The impact of cholesterol-lowering prescription drugs on GC risk was evaluated, using the “pROC” package in R (v 1.18.0) (8) to plot ROC curves.

### 2.7 Statistical analysis

Bioinformatics analyses were performed utilizing the R programming language (v 4.2.2). Unless otherwise specified, p < 0.05 was considered statistically significant.

## 3 Results

### 3.1 Baseline statistical description

Baseline statistics showed that the use of cholesterol-lowering prescription medications showed a significant intergroup difference with GC (p < 0.05). In addition, except for gender, race, BMI, smoke and SII, other covariates showed significant intergroup differences with GC (p < 0.05).(**Table 1**)

**Table 1.** Characteristics of NHANES participants.

| Table 1 Characteristics of NHANES participants |  |  |  |  |
| --- | --- | --- | --- | --- |
|  | level | Normal | GC | p |
| n |  | 51630 |  | 36 |
| Age (%) | Above_or_65 | 10544 (20.4) | 19 (52.8) | <0.001 |
|  | Below_65 | 41086 (79.6) | 17 (47.2) |  |
| Gender (%) | Male | 24909 (48.2) | 18 (50.0) | 0.965 |
|  | Female | 26721 (51.8) | 18 (50.0) |  |
| Race (%) | Mexican American | 9804 (19.0) | 9 (25.0) | 0.079 |
|  | Other Hispanic | 4032 ( 7.8) | 1 ( 2.8) |  |
|  | Non-Hispanic White | 22921 (44.4) | 10 (27.8) |  |
|  | Non-Hispanic Black | 10584 (20.5) | 13 (36.1) |  |
|  | Other Race | 4289 ( 8.3) | 3 ( 8.3) |  |
| Education (%) | Less Than High School | 14070 (27.3) | 17 (47.2) | 0.013 |
|  | High School graduate | 11974 (23.2) | 9 (25.0) |  |
|  | More Than High School | 25586 (49.6) | 10 (27.8) |  |
| PIR(%) | Low | 15960 (30.9) | 14 (38.9) | 0.034 |
|  | Middle | 19579 (37.9) | 18 (50.0) |  |
|  | High | 16091 (31.2) | 4 (11.1) |  |
| BMI(%) | Underweight | 818 ( 1.6) | 2 ( 5.6) | 0.09 |
|  | Normal | 14674 (28.4) | 14 (38.9) |  |
|  | Overweight | 17538 (34.0) | 8 (22.2) |  |
|  | Obese | 18600 (36.0) | 12 (33.3) |  |
| Smoke (%) | No | 28286 (54.8) | 14 (38.9) | 0.08 |
|  | Yes | 23344 (45.2) | 22 (61.1) |  |
| Hypertension (%) | No | 35184 (68.1) | 18 (50.0) | 0.031 |
|  | Yes | 16446 (31.9) | 18 (50.0) |  |
| Diabetes (%) | No | 45123 (87.4) | 27 (75.0) | 0.018 |
|  | Yes | 5578 (10.8) | 9 (25.0) |  |
|  | Borderline | 929 ( 1.8) | 0 ( 0.0) |  |
| SII (mean (SD)) |  | 565.30 (379.37) | 493.28 (333.43) | 0.255 |
| Medication for cholesterol (mean (SD)) |  | 0.01 (0.12) | 0.06 (0.33) | 0.013 |

### 3.2 Risk correlation analysis of medication for cholesterol and GC

Then, three consecutive multivariate glm regression models were constructed. The results indicated that the effect of cholesterol-lowering prescription medications on GC was not significantly confounded by other covariates (model 1: OR = 1.80, 95% CI = 1.23-2.63, p < 0.05; model 2: OR = 1.79, 95% CI = 1.13-2.83, p < 0.05; model 3: OR = 1.66, 95% CI = 1.03-2.68, p < 0.05).(**Table 2**)

**Table 2.** Risk correlation analysis.

| Table 2 Risk correlation analysis |  |  |  |
| --- | --- | --- | --- |
| exposure | model1_OR(95%_CI) | model2_OR(95%_CI) | model3_OR(95%_CI) |
| Medication for cholesterol | 1.8(1.23-2.63) | 1.79(1.13-2.83) | 1.66(1.03-2.68) |
| p_value | 0.00276 | 0.0134 | 0.0363 |

### 3.3 Significant correlation between medication for cholesterol and GC based on risk stratification

To further confirm the stability of the correlation between the use of cholesterol-lowering prescription medications and GC risk in different populations, a risk stratification analysis was conducted. The findings indicated that the two remained strongly correlated, with the use of cholesterol-lowering prescription medications being a risk factor for GC (OR = 1.73, 95% CI = 1.07-2.80, p < 0.05) (**Figure 2**).

**Figure 2.**
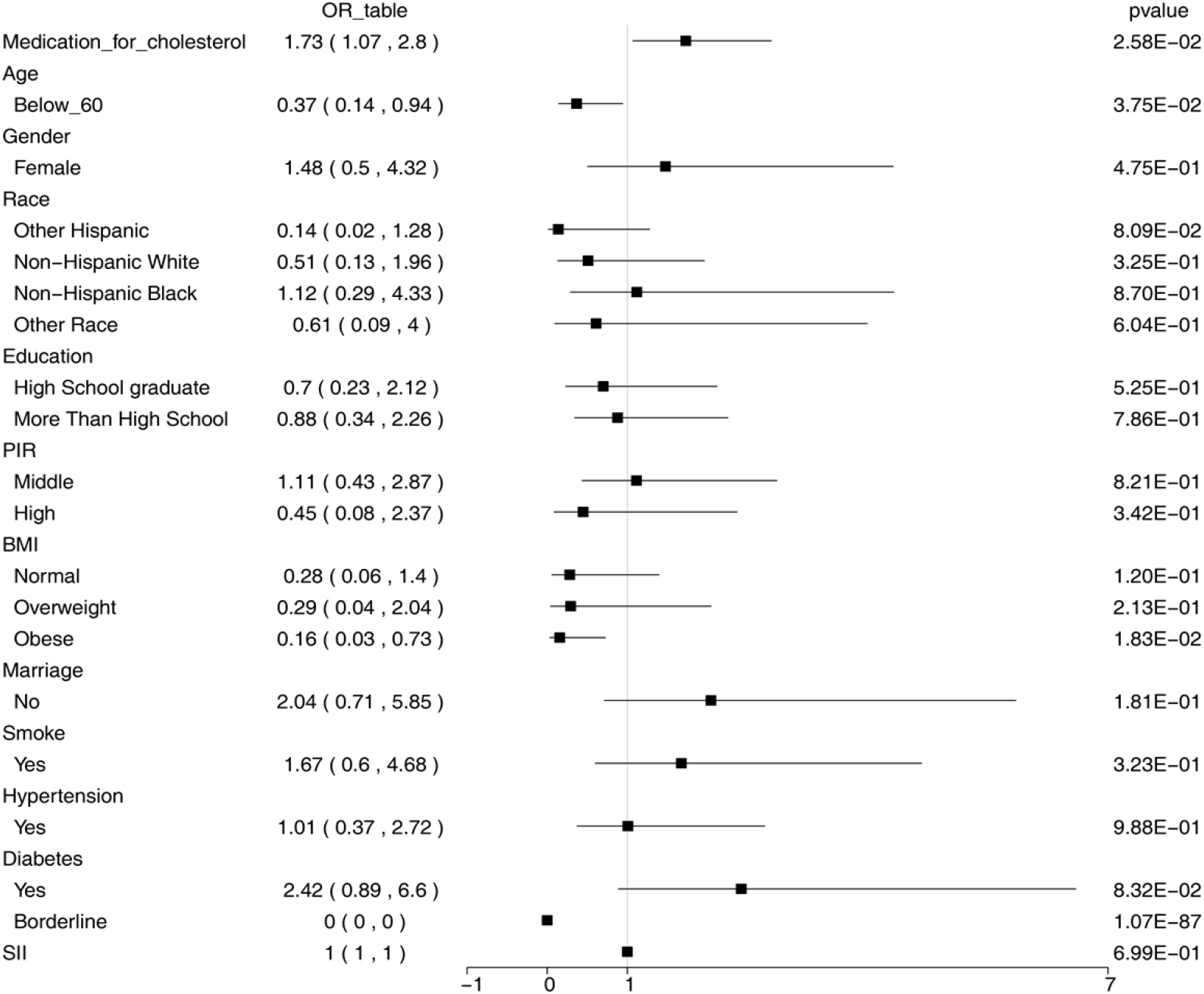
Stratified analysis of GC risk. Odds ratios (ORs) with 95% confidence intervals (CIs) were calculated across different demographic and clinical subgroups.

### 3.4 Evaluation of the impact of medication for cholesterol on GC based on ROC

The ROC analysis results showed that the use of cholesterol-lowering prescription medications had an AUC of 0.799 for predicting GC risk, indicating that the model had high accuracy in predicting GC risk factors (**Figure 3**).

**Figure 3.**
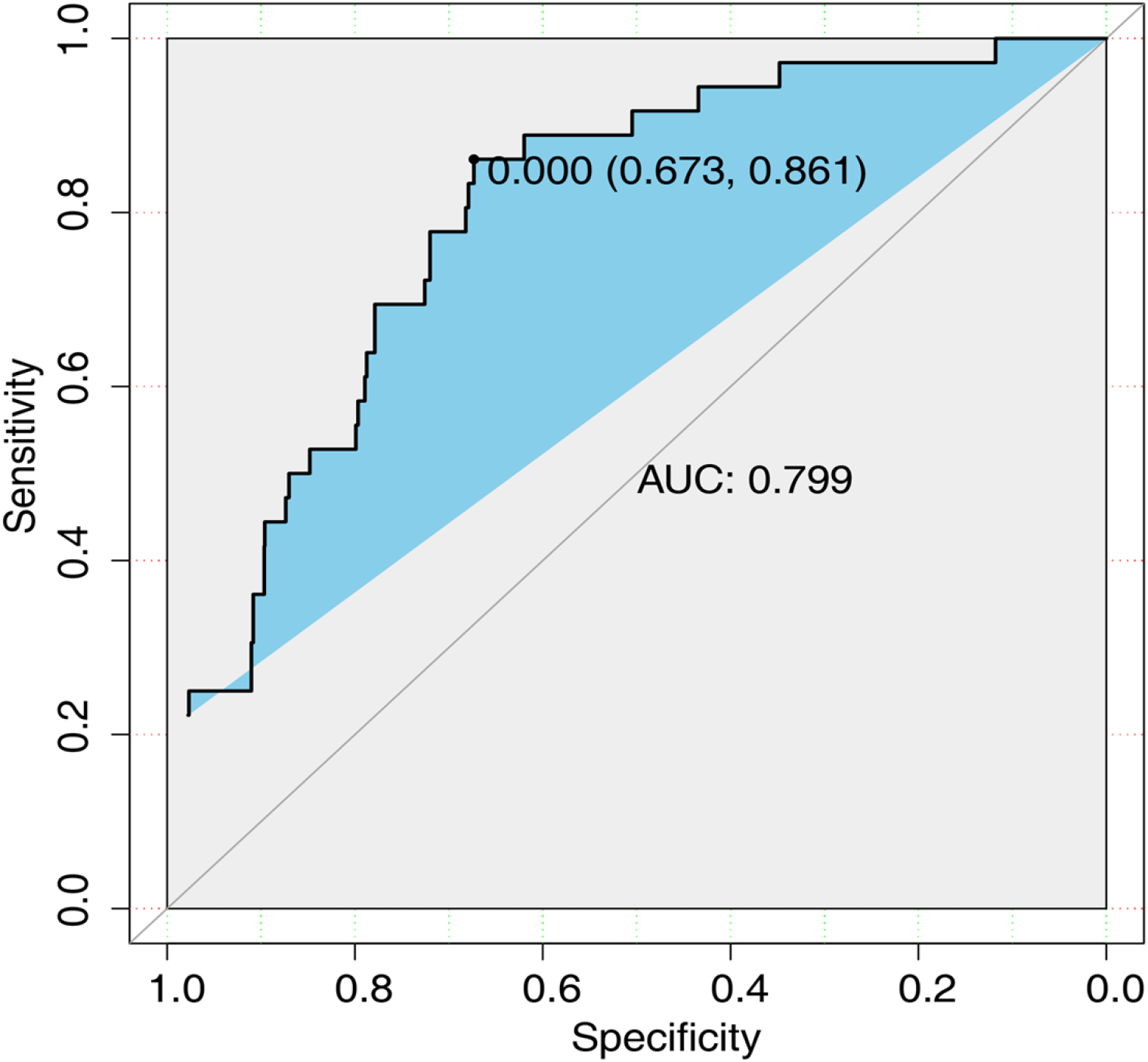
Receiver operating characteristic (ROC) curve analysis of cholesterol-lowering medication use for predicting GC risk. X-axis (1−Specificity/False Positive Rate): Indicates the proportion of individuals without gastric cancer who are incorrectly predicted as positive by the model. Y-axis (Sensitivity/True Positive Rate): Represents the proportion of individuals who actually have gastric cancer and are correctly predicted as positive by the model.

## 4 Discussion

GC is a significant contributor to the global cancer burden and is one of the most common and deadly malignancies worldwide (9). Cholesterol intake has been shown to significantly increase the risk of GC, and dysregulation of cholesterol metabolism may promote the development and progression of GC. However, despite the widespread use of cholesterol-lowering medications, their relationship with GC risk remains unclear. A systematic review and meta-analysis of observational studies demonstrated that elevated dietary cholesterol intake is significantly associated with a 35% increased risk of GC in adult populations(10). This study utilizes NHANES data to explore the potential association between cholesterol-lowering drugs and GC, aiming to provide evidence for personalized prevention and management strategies.

The present study reveals a significant positive association between the use of cholesterol-lowering medications and the risk of gastric cancer (odds ratio [OR] > 1.73, p < 0.05). This finding may be attributed to the pharmacological mechanisms of cholesterol-lowering drugs and their potential effects on the gastrointestinal microenvironment. For example,A large database analysis showed a widely used class of cholesterol-lowering agents may indirectly elevate gastric cancer risk by modulating cholesterol metabolism, inflammatory pathways, or cellular proliferation signaling cascades (11). Cholesterol, as a critical component of cell membranes, plays a vital role in maintaining cellular homeostasis. Dysregulation of cholesterol metabolism may disrupt the balance between cell proliferation and apoptosis, thereby fostering tumorigenesis. Furthermore, cholesterol-lowering drugs could influence gastric cancer risk by altering the composition of the gut microbiota or the local immune microenvironment (12) (13). However, these proposed mechanisms necessitate further validation through rigorous experimental studies and long-term epidemiological investigations.

The findings of this study exhibit certain discrepancies with other research investigating the relationship between medication use and gastric cancer. For instance, Liang Wang et al. demonstrated that molecular targeted agents (MTAs) in patients with advanced/metastatic gastric cancer may lead to severe adverse events, such as infections, gastrointestinal bleeding, and arterial thromboembolism (14). While MTAs primarily function by inhibiting tumor growth signaling pathways, their side effects may exacerbate risks in gastric cancer patients. In contrast, cholesterol-lowering drugs may influence gastric cancer development through metabolic or inflammatory mechanisms. On the other hand, Jeeyun Lee et al. reported that statin use in diabetic patients was associated with a reduced risk of gastric adenocarcinoma (15), which contrasts with the findings of the present study. This suggests that the relationship between cholesterol-lowering drugs and gastric cancer may vary depending on population characteristics (e.g., presence of diabetes) or drug type. Additionally, K.E.L. McColl highlighted that acid-suppressive medications, such as proton pump inhibitors, are associated with the prevalence of gastric and esophageal cancers (16). This may be attributed to long-term acid suppression leading to alterations in the gastric environment, such as reduced gastric acidity and bacterial overgrowth. Similarly, cholesterol-lowering drugs may increase gastric cancer risk by modifying the gastrointestinal environment or metabolic pathways. Collectively, these studies indicate that the relationship between medication use and gastric cancer is complex and multifaceted, likely influenced by a combination of drug type, mechanism of action, and population characteristics.

The results demonstrated a significant association between cholesterol-lowering medications and GC. This association remained robust as an independent exposure factor and after adjustment for multiple covariates, suggesting a potential role of these medications in promoting GC development. Stratified risk analysis further supported this positive association (OR=1.73, 95% CI=1.07-2.8, p 0.05). These findings highlight the need for closer gastric health monitoring among long-term users of cholesterol-lowering drugs, especially those with other GC risk factors such as Helicobacter pylori infection or a family history of GC. Moreover, receiver operating characteristic (ROC) analysis showed that the use of cholesterol-lowering medications held some predictive value for GC, with an area under the curve (AUC) of 0.799. However, its diagnostic accuracy as a single marker was restricted. Future research should consider integrating this factor with other biomarkers or risk factors. A systematic review and meta-analysis retrieved articles from PubMed, Scopus, and Google Scholar up to May 2021, This systematic review specifically investigated the association between dietary cholesterol intake and GC risk. Summary effect estimates were derived through random-effects meta-analysis, with pooled odds ratios (ORs) and corresponding 95% confidence intervals (CIs) calculated to quantify the magnitude of association The meta-analysis revealed a significant positive association between elevated dietary cholesterol intake and GC risk, with adults in the highest consumption quartile demonstrating a 35% increased risk of GC development compared to those in the lowest intake group (11).

These findings emphasize the importance of dietary cholesterol regulation in GC prevention. Contrastingly, a review by Bai et al. suggested a negative correlation between the intake of statins or metformin and GC risk, potentially due to mechanisms such as apoptosis induction and anti-angiogenesis effects of statins (17). A meta-analysis supported a protective effect of statins against GC. However, observational bias and inconsistent findings from randomized controlled trials (RCTs) precluded recommending statins for GC prevention (18). Another meta-analysis, encompassing Asian and Western populations (94.8% of GC cases, n=5290), showed a non-significant chemopreventive effect of statins (adjusted OR=0.83; 95% CI=0.66-1.05). Despite a statistical trend, the included RCTs lacked sufficient power to detect meaningful differences in GC incidence (19).

## 5 Conclusion

In summary, this study identified a significant association between the use of cholesterol-lowering prescription medications and increased GC risk, suggesting that these medications may influence GC development. These findings offer valuable guidance for future research and provide insights into GC screening strategies for high-risk populations.

## 7 Data Availability Statement

The datasets for this study can be found in the NHANES database (https://www.cdc.gov/nchs/nhanes/index.htm).

## 8 Human Ethics and Consent to Participate declarations

not applicable.

## 9 Acknowledgments

The authors are thankful to the contributors of this Research Topic, as well as to the editorial support provided by the Journal.

## 10 Clinical trial number: not applicable

## 11 Every human participant provide their consent

## 12 there was no Funding

